# Investigating the relationships between unfavorable sleep and metabolomic traits: evidence from multi-cohort multivariable regression and Mendelian randomization analyses

**DOI:** 10.1101/2020.08.27.20173518

**Authors:** Maxime M Bos, Neil J Goulding, Matthew A Lee, Amy Hofman, Mariska Bot, René Pool, Lisanne S Vijfhuizen, Xiang Zhang, Chihua Li, Rima Mustafa, Matt J Neville, Ruifang Li-Gao, Stella Trompet, Marian Beekman, Nienke R Biermasz, Dorret I Boomsma, Irene de Boer, Constantinos Christodoulides, Abbas Dehghan, Ko Willems van Dijk, Ian Ford, He Gao, Mohsen Ghanbari, Bastiaan T Heijmans, M Arfan Ikram, J Wouter Jukema, Dennis O Mook-Kanamori, Fredrik Karpe, Annemarie I Luik, L.H. Lumey, Arn M.J.M. van den Maagdenberg, Simon P Mooijaart, Renée de Mutsert, Brenda W.J.H. Penninx, Patrick CN Rensen, Rebecca C Richmond, Frits R Rosendaal, Naveed Sattar, Robert A Schoevers, P Eline Slagboom, Gisela M Terwindt, Carisha S Thesing, Kaitlin H Wade, Carolien A Wijsman, Gonneke Willemsen, Aeilko H. Zwinderman, Diana van Heemst, Raymond Noordam, Deborah A Lawlor

## Abstract

**Background:** Sleep traits are associated with cardiometabolic disease risk, with evidence from Mendelian randomization (MR) suggesting that insomnia symptoms and shorter sleep duration increase coronary artery disease risk. We combined adjusted multivariable regression (AMV) and MR analyses of phenotypes of unfavourable sleep on 113 metabolomic traits to investigate possible biochemical mechanisms linking sleep to cardiovascular disease.

**Methods:** We used AMV (N=17,370) combined with two-sample MR (N=38,618) to examine effects of self-reported insomnia symptoms, total habitual sleep duration, and chronotype on 113 metabolomic traits. The AMV analyses were conducted on data from 10 cohorts of mostly Europeans, adjusted for age, sex and body mass index. For the MR analyses, we used summary results from published European-ancestry genome-wide association studies of self-reported sleep traits and of nuclear magnetic resonance (NMR) serum metabolites. We used the inverse-variance weighted (IVW) method and complemented this with sensitivity analyses to assess MR assumptions.

**Results:** We found consistent evidence from AMV and MR analyses for associations of usual vs. sometimes/rare/never insomnia symptoms with lower citrate (−0.08 standard deviation (SD)[95% confidence interval (CI): −0.12, −0.03] in AMV and −0.03SD [−0.07, −0.003] in MR), higher glycoprotein acetyls (0.08SD [95%CI: 0.03, 0.12] in AMV and 0.06SD [0.03, 0.10) in MR]), lower total very large HDL particles (−0.04SD [−0.08, 0.00] in AMV and - 0.05SD [−0.09, −0.02] in MR) and lower phospholipids in very large HDL particles (−0.04SD [−0.08, 0.002] in AMV and −0.05SD [−0.08, −0.02] in MR). Longer total sleep duration associated with higher creatinine concentrations using both methods (0.02SD per 1-hour [0.01, 0.03] in AMV and 0.15SD [0.02, 0.29] in MR) and with isoleucine in MR analyses (0.22SD [0.08, 0.35]). No consistent evidence was observed for effects of chronotype on metabolomic measures.

**Conclusions:** Whilst our results suggested that unfavourable sleep traits may not cause widespread metabolic disruption, some notable effects were observed. The evidence for possible effects of insomnia symptoms on glycoprotein acetyls and citrate and longer total sleep duration on creatinine and isoleucine might explain some of the effects, found in MR analyses of these sleep traits on coronary heart disease, which warrant further investigation.

## Introduction

Several systematic reviews and large biobank studies have reported associations of self-reported insomnia symptoms, short and long sleep duration, and chronotype (i.e., having an evening rather than morning preference) with increased risk of cardiovascular disease, type 2 diabetes and risk factors for these^1-9^. The mechanisms underlying these associations are unclear, and it is plausible that specific sleep traits may contribute to the misalignment of various behavioral and internal physiological processes, including aspects of metabolism that causes adverse cardiometabolic health.

There is some evidence of poor sleep quality, shorter sleep duration and having an evening chronotype being associated with higher triglyceride, total cholesterol and low-density lipoprotein cholesterol (LDL-C) levels and lower high-density lipoprotein cholesterol (HDL-C) concentrations^10-12^. However, the extent to which these associations are explained by confounding factors, such as body mass index^11^, is unclear. Beyond conventional multivariable-adjusted regression analyses, we have previously demonstrated that sleep duration modifies the associations of genetic variation with triglycerides, LDL-C and HDL-C in a large sleep-gene interaction analysis, suggesting that possible different biological mechanisms underlie the associations of short and long sleep duration with these lipid traits^13^. However, these genetic interaction analyses do not assess causality and, like previous multivariable-adjusted regression analyses, have focused on a limited number of lipid traits.

Mendelian randomization (MR) uses genetic variants that are robustly associate with an exposure as an instrumental variable to obtain unconfounded effects of that exposure on an outcome of interest^14-16^. Recent MR analyses have suggested a causal effect of insomnia symptoms on coronary heart disease^17^ and of short (<6 hours) sleep duration on myocardial infarction risk^18^.

The aim of this study was to determine the possible causal effect of sleep traits on metabolomic traits. We compared findings from adjusted multivariable regression (AMV) and MR analysis, to determine the relationships between self-reported insomnia symptoms (usually vs sometimes/rare/never), total habitual sleep duration (per 1 hour longer) and chronotype (evening vs morning preference) and 113 nuclear magnetic resonance (NMR) metabolomic traits. Cross-sectional AMV was performed with adjustment for age, sex and BMI in 17,370 individuals from 10 cohorts of mostly Europeans. Two-sample MR used summary results from genome-wide association studies (GWAS) of different sleep traits in 1,331,010 (insomnia)^19^, 446,118 (sleep duration)^20^, and 651,295 (chronotype)^21^ European adults and summary results from four GWAS of 113 circulating metabolomic measures from NMR in 38,618 European adults. In secondary analyses, we explored effects of short (<7 vs 7-<9 hours) and long (≥9 vs 7-<9 hours) sleep duration on the metabolomic traits. We highlight results that were consistent across both methods, as the different key sources of bias of the two methods (e.g., residual confounding in AMV and unbalanced horizontal pleiotropy in MR, respectively) mean that, where there is consistency, this is more likely to reflect a causal effect^22^.

## Methods

### Studies used for AMV

Cross-sectional AMV analyses were performed using data from 10 cohorts: the Active and Healthy Ageing (AGO) study^23^, the Dutch Hunger Winter Families Study (DWFS)^24^, the Healthy Life in an Urban Setting (HELIUS) Study^25^, the Leiden University Migraine Neuro-Analysis (LUMINA)^26^, Netherlands Study of Depression and Anxiety (NESDA)^27^, the Netherlands Twin Register (NTR)^28^, the Netherlands Epidemiology of Obesity (NEO)

Study^29^, and the Rotterdam Study cohorts 1, 2 and 3 (RS1, RS2 and RS3)^30^. Details, and distributions for characteristics, of each study are given in the **Supplemental Material** and **Supplemental Table S1**. Each participating study obtained written informed consent from all participants and received approval from the appropriate local institutional review boards. Before the analyses, we excluded all participants with diabetes (defined as self-report/hospital record, fasting plasma glucose >7 mmol/L and/or use of hypoglycaemic medication) given the known disturbances on many metabolomic traits.

### Studies used for MR analyses

We performed two-sample MR analyses using publicly available summary-level data^14^ from the following GWAS:

### Sleep trait GWAS

We selected genome-wide significant (p-value<5e-8) variants as instrumental variables from the following GWAS:

- **Insomnia**: A GWAS that pooled data from two large biobanks (UK Biobank and 23andMe) and included 1,331,010 unrelated European-ancestry adults. This GWAS identified 248 variants for experience of insomnia symptoms (usually vs sometimes/rare/never)^19^.
- **Sleep duration**: A GWAS undertaken in UK Biobank of 446,118 unrelated European-ancestry adults^20^. This GWAS identified 78 variants for total sleep duration (mean 7.2h; SD1.1h). In addition, this GWAS identified 27 variants for short sleep duration (<7 h vs 7 to <9 hours; N = 106,192 cases) and 8 for long sleep duration (≥9 h vs 7 to <9 hours; N = 34,184 cases).
- **Chronotype**: A GWAS that pooled data from two large biobanks (UK Biobank and 23andMe) and included 697,828 unrelated European-ancestry adults (651,295 of whom were in the combined (both biobanks) GWAS of morning versus evening preference that we have used in this two-sample MR study. This GWAS identified 351 variants for chronotype^21^. Because previous observational studies have found increased risk of cardiometabolic diseases and risk factors in those with an evening preference, we transformed the GWAS results to reflect alleles associated with evening preference.

### NMR Metabolite GWAS

- MAGNETIC consortium (N = 24,925)^31^ with summary-level GWAS data downloaded from http://www.computationalmedicine.fi/data#NMRGWAS
- In addition, to increase statistical power in the MR analyses, we generated new summary-level GWAS data from three cohorts using similar analyses procedures to the MAGNETIC consortium:

- Oxford Biobank (N = 6,616)^32^;
- NEO (N = 4,734)^29^;
- Pravastatin in Elderly Individuals at Risk of Vascular Disease (PROSPER) (N = 2,343; placebo arm only)^33^.

All GWASs were undertaken in participants of European ancestry. Further details of the MAGNETIC consortium and the three additional cohorts are provided in **Supplemental Methods**. There was no overlap between the cohorts included in the sleep trait GWAS and those included in the NMR GWAS.

### Sleep traits

In both AMV and MR analyses, sleep traits were self-reported and analysed in the same units/categories. Insomnia symptoms were assessed with a question similar to “Do you have trouble falling asleep at night or do you wake up in the middle of the night?” with the following answers possible: “never/rarely”, “sometimes”, “usually”, or “prefer not to answer”. In the AMV and GWAS analyses participants who answered “usually” were defined as having insomnia symptoms and were compared to those answering “never/rarely” or “sometimes”. Habitual sleep duration was assessed using a question similar to “On an average day, how many hours of sleep do you get?”. For our main analyses, we examined effects of total sleep duration (per 1 hour longer) on metabolomic measures. In secondary analyses, we explored associations of short (<7 vs 7-<9) and long (≥9 vs 7-<9 hours) habitual sleep. These latter two analyses were considered exploratory because of lower statistical power and possible weak instrument bias in the MR analyses. For chronotype, a question similar to “Are you naturally a night person or a morning person?” with the possible responses “Night owl/night person”, “Early bird/morning person”, “Neither/not sure” was used in most studies. A variation on the question in UK Biobank included more responses: “Definitely a morning person”, “More a morning than evening person”, “More an evening than a morning person”, “Definitely an evening person”, “Do not know”. Participants were classified as having a ‘morning preference’ (“Early bird/morning person”, “Definitely a morning person” or “More a morning than evening person”), the reference group, or an ‘evening preference’ (“Night owl/night person”, “More an evening than a morning person” or “Definitely an evening person”). For all traits those responding “do not know”, “unsure” or “prefer not to answer” were excluded.

### NMR-based metabolomic profiling

In both the metabolite GWAS and studies included in the AMV meta-analysis, metabolites were quantified using a high-throughput proton (^1^H) NMR metabolomics platform^34^ (https://nightingalehealth.com/) to quantify a maximum of 148 (excluding ratios) lipid and lipoprotein and metabolite concentrations in fasting serum or plasma samples. The quantitative NMR measures include numerous lipid species and fatty acids, as well as some amino acids, markers of glucose homeostasis, fluid balance and an inflammatory marker. This platform has been used widely in population-based studies of cardiometabolic diseases, and has been described in detail elsewhere^34-36^. There were 113 metabolomic trait measurements that were available for both AMV and MR analyses.

### Statistical Analyses

In both AMV and MR analyses, we estimated the same effect: the difference in mean NMR metabolites (SD units of the natural log-transformed metabolomic traits; as dependent variables) comparing (i) usually experiencing insomnia symptoms to sometimes, rarely or never, (ii) per 1 hour longer habitual sleep duration and (iii) an evening to a morning preference. All analyses were performed in R (v3.6.1)^37^.

### Multivariable-adjusted regression meta-analysis

Cross-sectional AMV was performed by each of the individual cohorts according to a pre-specified analysis plan and standardized analysis script. Results were collected centrally for quality control subsequent fixed-effect meta-analyses using the R “rmeta” package, using similar procedures as described previously^38^. AMV analyses adjusted for age, sex and BMI.

### MR analyses

We excluded all palindromic single nucleotide polymorphisms (SNPs) and those in linkage disequilibrium at R^2^>0.001 (based on the 1000genomes (phase 1) panel). After these exclusions, we searched for all remaining independent sleep-associated variants (149 for insomnia, 57 for total sleep duration [and an additional 25 and 71 variants for short and long habitual sleep duration, respectively] and 208 for chronotype) in the GWAS of NMR metabolomic measures, and the directions of the summary data were harmonised (i.e. making sure that each effect estimate was coded in the same direction with respect to the effect allele as SNP associations from the summary sleep trait data) with those of the sleep trait summary data.

The MRCIEU/TwoSampleMR package was used for harmonization of the exposure and outcome SNPs and to perform the MR analyses^16^. For our main analyses, we used the multiplicative random effects inverse variance-weighted (IVW) approach^39^. This method generates a causal estimate of the sleep traits on metabolomic traits by regressing the SNP-sleep trait association on the SNP-metabolomic measure association, weighted by the inverse of the SNP-metabolomic measure association, and constraining the intercept of this regression to zero. Standard errors are corrected to take into account any between SNP heterogeneity and assumes that there is no directional horizontal pleiotropy. To explore this assumption further, we performed sensitivity analyses using MR-Egger^40^ and weighted-median estimator^41^ methods. MR-Egger is similar to the IVW method but does not force the regression line (i.e., of the SNP-sleep trait association on the SNP-metabolomic measure association) through an intercept of zero. It is statistically less efficient (providing wider confidence intervals) but provides a causal estimate (i.e., the regression slope) that is corrected for directional horizontal pleiotropy, and a non-zero intercept is an indication of the existence of directional pleiotropy. The weighted-median estimator is valid if more than 50% of the weight of the genetic instrument is from valid variants (i.e., if one single SNP or several SNPs jointly contributing 50% or more of the weight in the MR analysis exhibit horizontal pleiotropy the calculated effect estimate may be biased).

We performed MR analyses for all sleep traits with each of the 4 metabolomic GWAS data sources (MAGNETIC, Oxford Biobank, NEO and PROSPER) and the results were subsequently meta-analysed using fixed-effect meta-analyses as implemented in the R package rmeta.

### Comparing multivariable regression and MR analysis results

Circos plots were used to summarise and visually compare the AMV and the IVW MR results. These were generated using R package EpiViz (version 0.1.0) (further details in the **Supplemental material**). We also generated scatter plots of the AMV vs MR results for each metabolite and compared the linear fit across all metabolites to a slope of perfect concordance and used R^2^ as a measure of goodness of fit (agreement) between the two methods across all 113 metabolomic traits.

Having compared results for the AMV and IVW MR methods across all metabolites, we then selected all sleep trait-metabolite associations that reached a pre-defined p-value threshold in AMV or IVW MR. We then compared results across AMV, IVW MR, MR-Egger and weighted median MR for those selected associations. Whilst we focus on results reaching a pre-defined p-value threshold in either AMV or IVW MR in the main paper and our conclusion, a full set of all results (AMV, unadjusted MV, IVW MR and all MR sensitivity analyses) are presented in Supplemental **Tables S2 to S11** in the **Supplemental Tables File**. We applied the same Bonferroni multiple testing corrected p-value threshold separately to the AMV and MR analyses. The threshold was determined taking into account the correlation structure of the metabolomic measures by using information from previous studies that have identified 17 principal components, which explain 95% of the metabolomic traits data variance^42^. Therefore, the two-sided threshold of *P <* 0.05 adjusted for multiple testing becomes *P <* 0.0029 (0.05/17). For any association that passed this threshold with either AMV or IVW MR we considered the result from the second method to be consistent if the point estimate had a similar direction of effect and the p-value for the second association was <0.05. This was justified on the basis that once one method passed the Bonferroni threshold, we were treating that result as a hypothesised effect and seeking replication and triangulation in the second method.

## Results

Full results of all AMV and MR analysis, including MR sensitivity analysis results, are presented in Supplemental **Tables S2 to S11** in the **Supplemental Tables File**.

### Insomnia symptoms

Visually inspecting the circos plot shows there was directional consistency between the AMV and IVW MR results for most of the metabolomic traits (**Figure 1**). With both methods, insomnia symptoms were associated with higher concentrations of small and medium very large density lipoprotein (VLDL) particles, small HDL particles and glycoprotein acetyls, and with lower concentrations of large HDL particles. Across all 113 metabolomic traits, there was good concordance of effect size and direction (**Figure 2**; R^2^ = 0.57).

**Figure 1:**
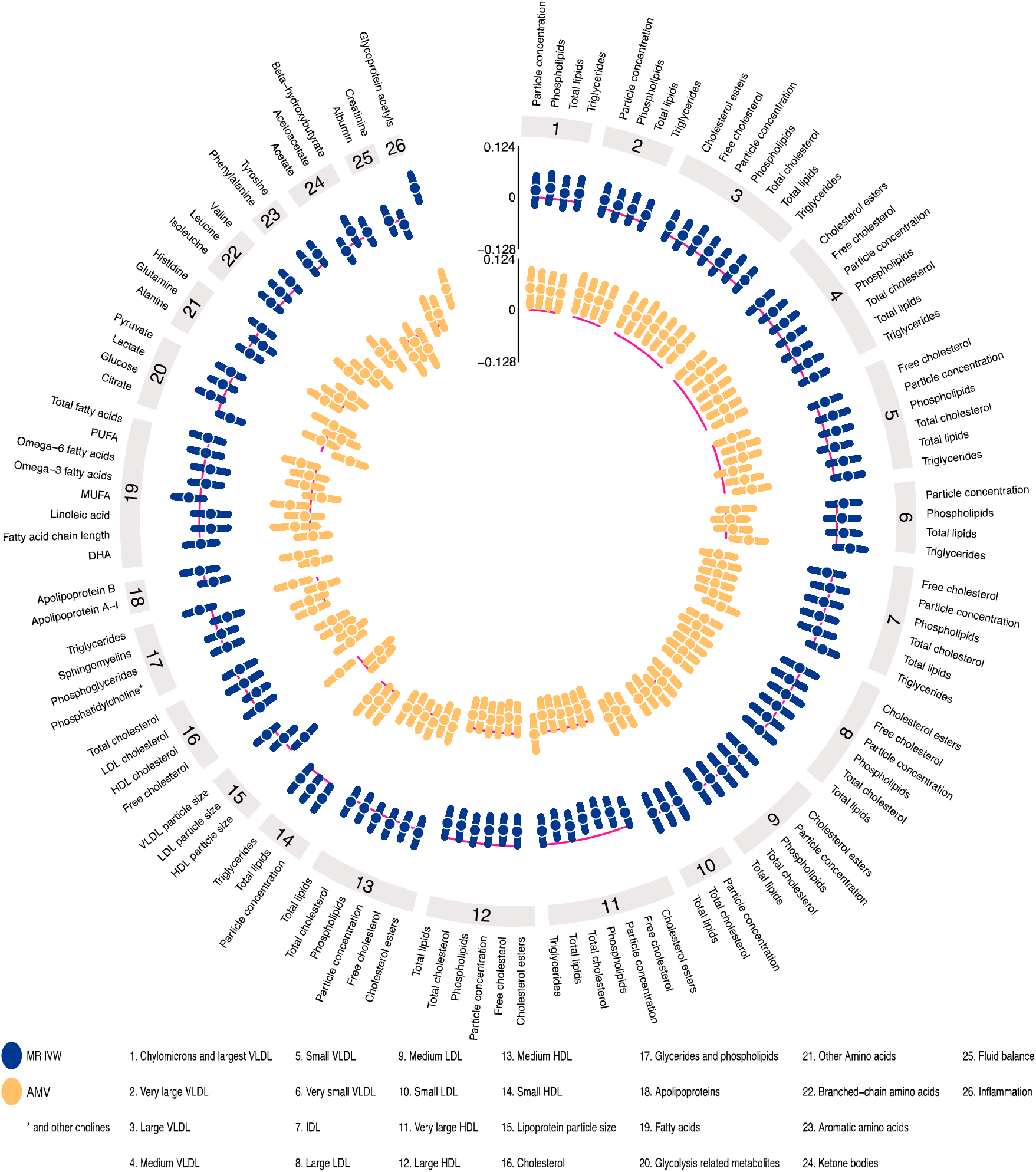
IVW Mendelian randomization estimates and age-, sex- and BMI-adjusted multivariable regression estimates for the associations between insomnia symptoms and 113 NMR-derived metabolomic measures. Results are expressed as the difference in mean metabolite concentrations (in standard deviation units) between those reporting usually versus sometimes/rarely/never experiencing insomnia symptoms. Abbreviations: AMV, adjusted (age, sex, BMI) multivariable regression; BMI, body mass index; IDL, intermediate density lipoprotein; IVW MR, Inverse variance weighted Mendelian randomization; LDL, low density lipoprotein; NMR, nuclear magnetic resonance; VLDL, very large density lipoprotein.

**Figure 2:**
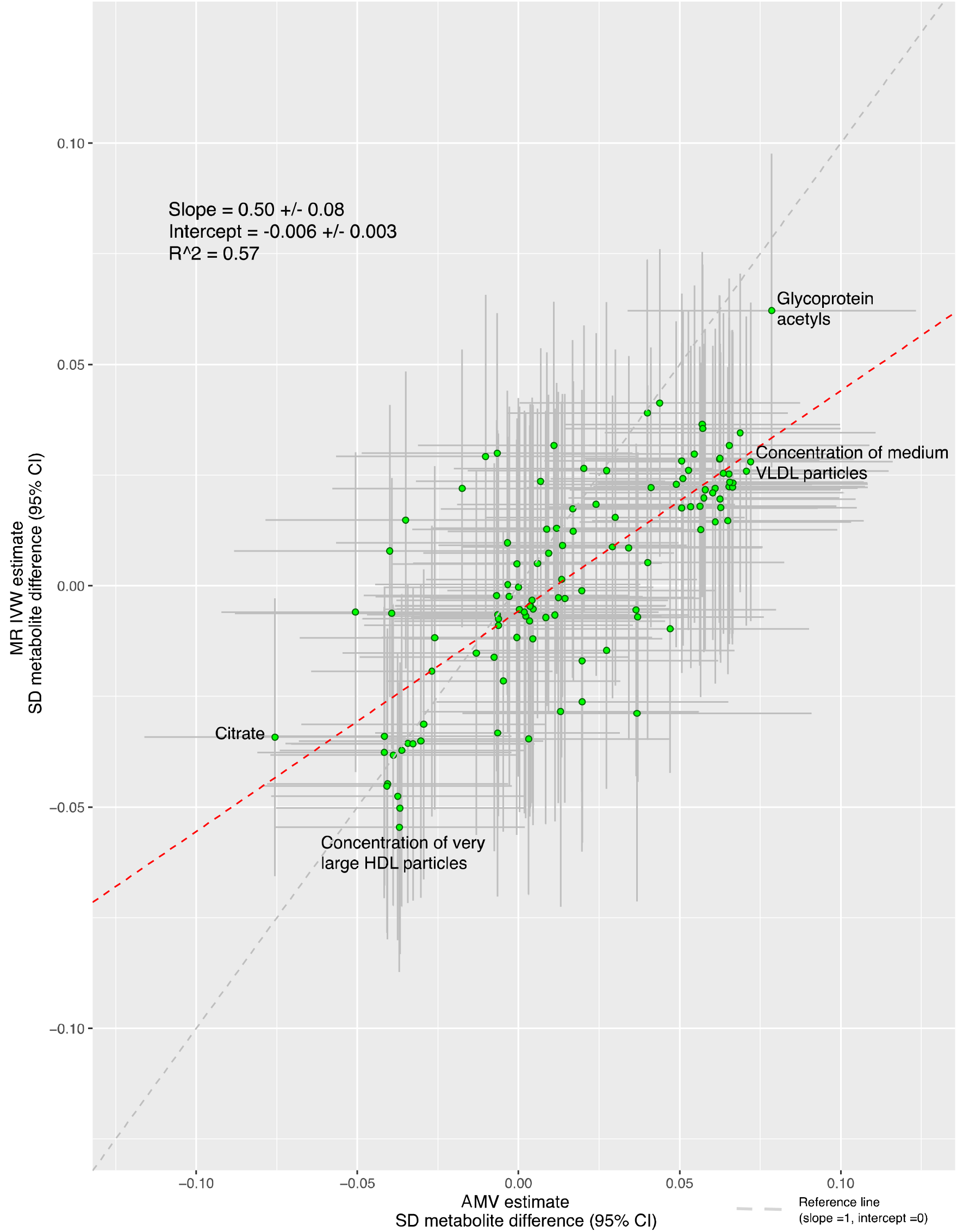
Comparison of the point estimates of the IVW Mendelian randomization and age-, sex- and BMI-adjusted multivariable regression analyses for the associations between insomnia symptoms and 113 NMR-derived metabolomic measures. Each green dot in the scatter plot represents a metabolic trait and the positions of the dots are determined by the differences in mean metabolite concentrations (in standard deviation units) between those reporting usually versus sometimes/rarely/never experiencing insomnia symptoms. These are estimated by Inverse variance weighted (IVW) Mendelian randomization (vertical axes) and age, sex and BMI adjusted multivariable regression (horizontal axes). The vertical grey lines for each dot indicate the 95% confidence intervals (CI) for the Mendelian randomization estimates and the horizontal grey lines for each dot indicate the 95% CI for the adjusted multivariable regression estimates. A linear fit (red dashed line) summarizes the similarity between the two estimates. A slope of 1 with an intercept of 0 (dashed grey line), with all green dots sitting on that line (R^2^ = 1), would indicate identical magnitude and direction between the two methods. R indicates goodness of linear fit and is a measure of the consistency between the two estimates. Abbreviations: AMV, adjusted (age, sex, BMI) multivariable regression; BMI, body mass index; CI, confidence interval; DHA, 22:6, docosahexaenoic acid; IVW MR, Inverse variance weighted Mendelian randomization, SD, standard deviation.

Associations of insomnia symptoms with the 113 metabolomic traits passed the multiple testing threshold *(P* < 0.0029) for 13 in the AMV analyses, and for 3 in the MR analyses (glycoprotein acetyls passed the threshold in both). Based on our pre-specified definition of consistency (i.e. same direction and p-value <0.05 in MR for any AMV results reaching the corrected p-value, and vice versa) we found consistent evidence from AMV and MR analyses for 4 associations. Specifically, usual vs. sometimes/rare/never insomnia symptoms lowered citrate (−0.08SD [95%CI: −0.12, −0.03] in AMV and −0.03SD [−0.07, - 0.003] in MR), increased glycoprotein acetyls (0.08SD [95%CI: 0.03, 0.12] in AMV and 0.06 [0.03, 0.10] in MR), and lowered total very large HDL particles (−0.04SD [−0.08, 0.00] in AMV and −0.05SD [−0.09, −0.02] in MR) and phospholipids in very large HDL particles (− 0.04SD [−0.08, 0.002] in AMV and −0.05SD [−0.08, −0.02] in MR) (**Figure 3**). MR sensitivity analyses were generally consistent with the main IVW analyses though confidence intervals were wide for the MR-Egger results. (**Figure 3**).

**Figure 3:**
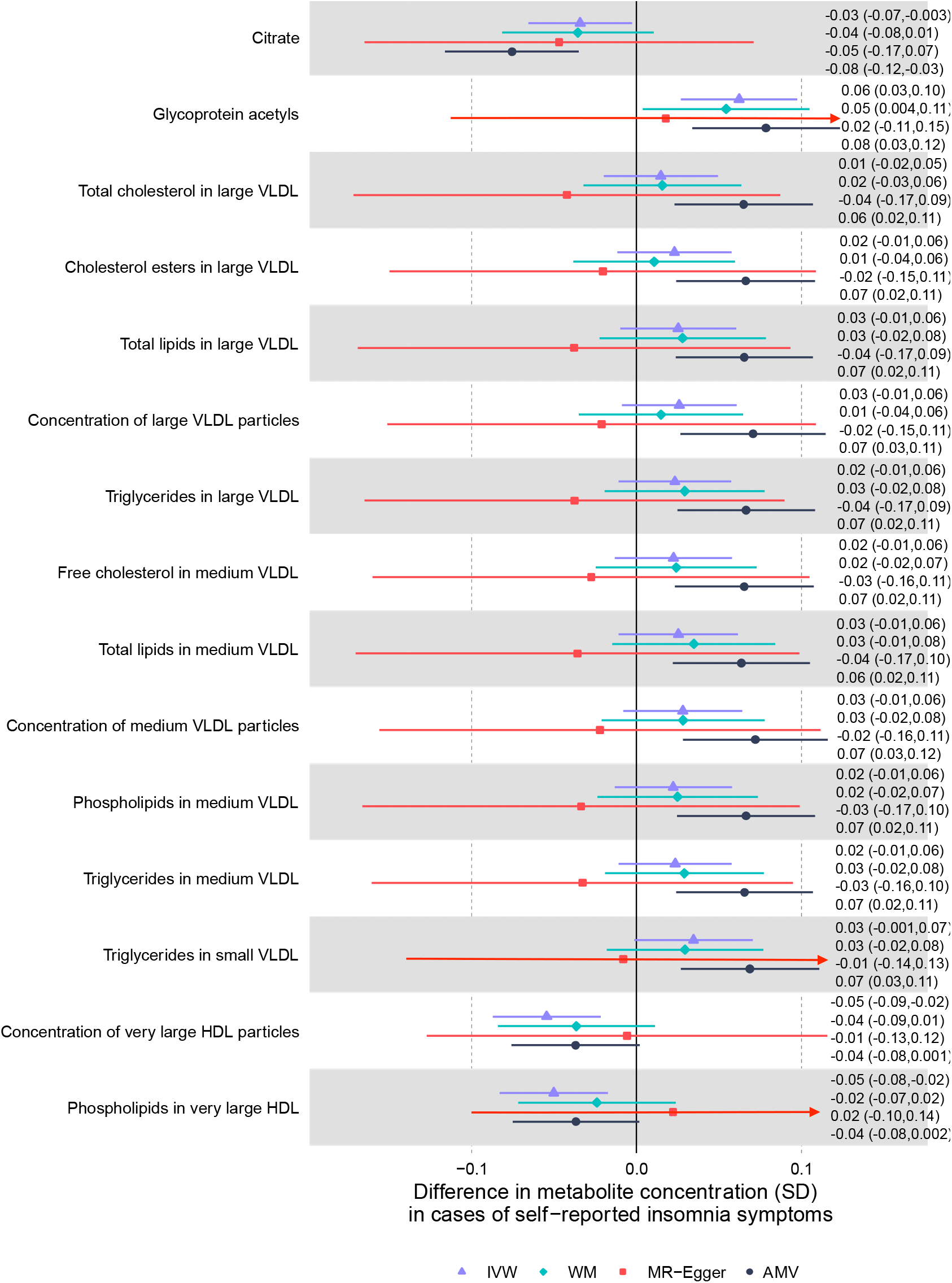
Mendelian randomization and age-, sex- and BMI-adjusted multivariable regression analyses results for select associations of insomnia symptoms with NMR- derived metabolomic measures. Figure shows inverse variance weighted (IVW) Mendelian randomization, Mendelian randomization sensitivity (weighted median (WM) and MR-Egger) and adjusted multivariable (AMV) regression analysis results. Results presented were selected on the basis of passing multiple testing threshold for either IVW or AMV (p-values < 0.0029) Theestimates are the difference in mean metabolite (in standard deviation units) between those reporting usually versus sometimes/rarely/never experiencing insomnia symptoms. Abbreviations: AMV, adjusted (age, sex, BMI) multivariable regression; BMI, body mass index; IVW MR, Inverse variance weighted Mendelian randomization; NMR, nuclear magnetic resonance; SD, Standard Error; VLDL, very low density lipoprotein; WM, Weighted Median.

### Sleep duration

Several associations of total sleep duration with metabolomics traits were directionally consistent in the the AMV and MR analyses (**Figure 4**). Where association directions were consistent, the MR results often had a stronger magnitude of association than the AMV results. Consistency of magnitude (as well as direction) was poor to moderate between the two methods (**Figure 5**, R^2^ = 0.37).

**Figure 4:**
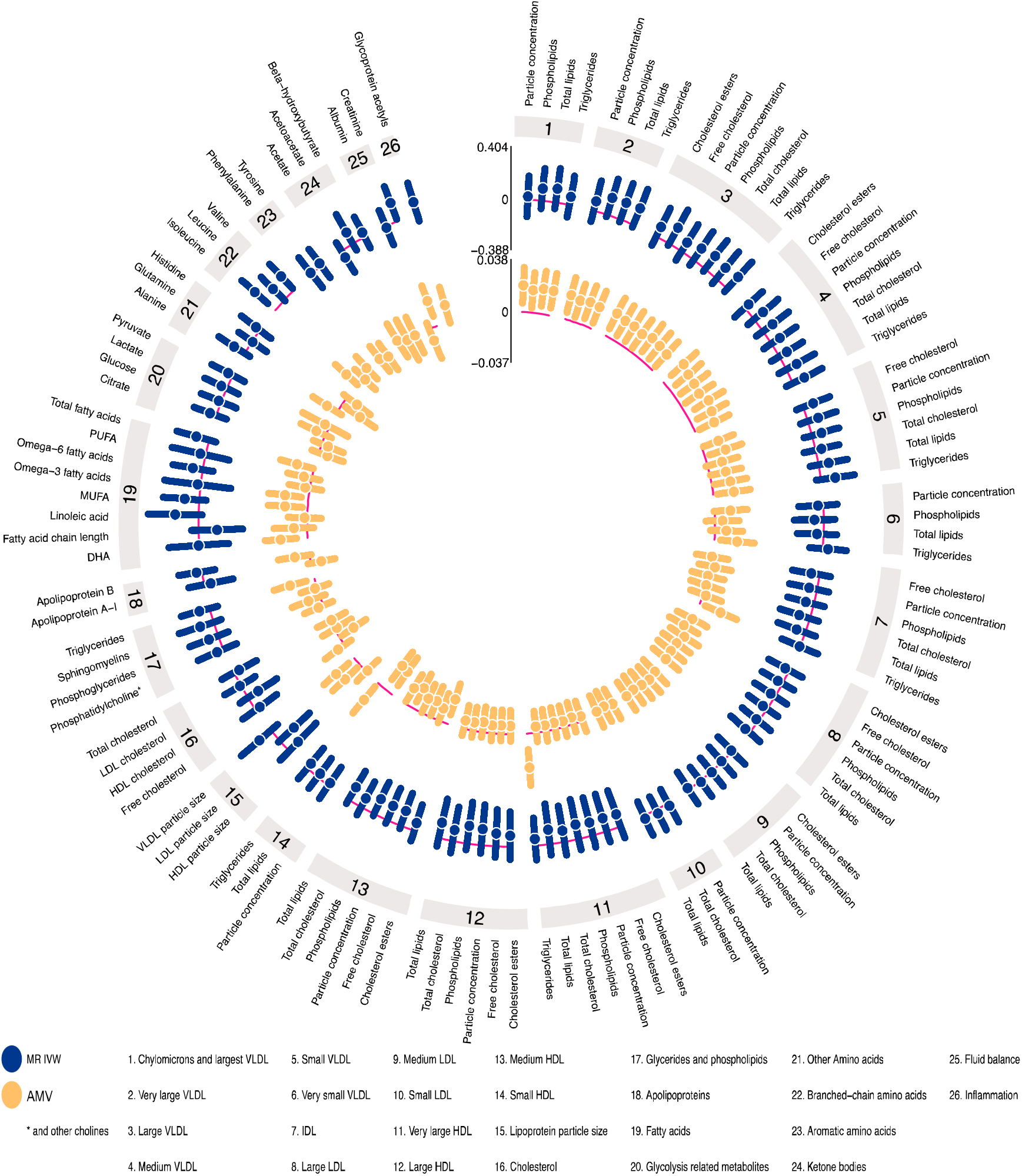
IVW Mendelian randomization estimates and age-, sex- and BMI-adjusted multivariable regression estimates for the associations between total sleep duration and 113 NMR-derived metabolomic measures. Results are expressed as the difference in mean metabolite concentrations (in standard deviation units) for each 1 hour greater reported total sleep duration. For visualization purposes the axes have unequal scaling. Abbreviations: AMV, adjusted (age, sex, BMI) multivariable regression; BMI, body mass index; IDL, intermediate density lipoprotein; IVW MR, Inverse variance weighted Mendelian randomization; LDL, low density lipoprotein; NMR, nuclear magnetic resonance; VLDL, very large density lipoprotein.

**Figure 5:**
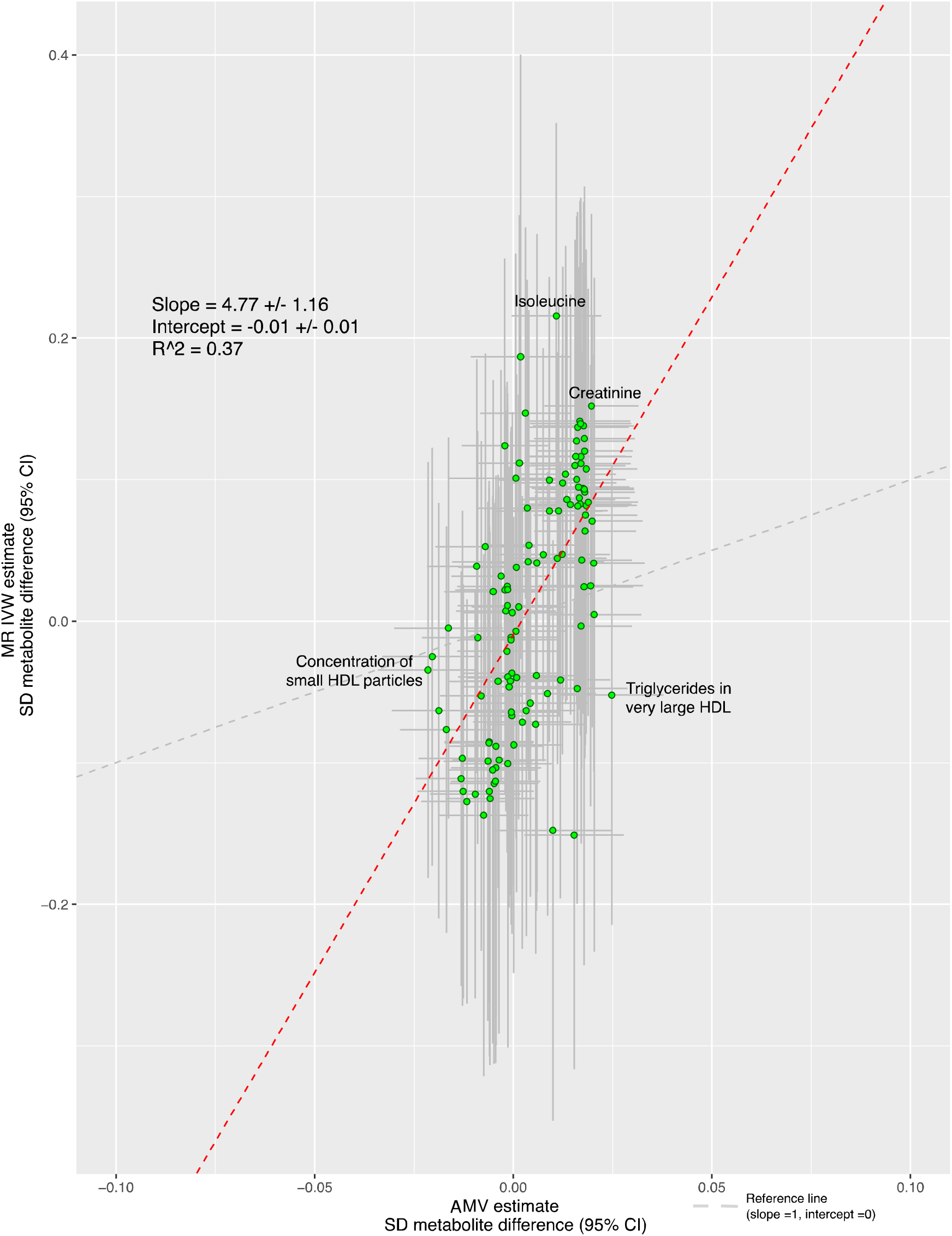
Comparison of the point estimates of the IVW Mendelian randomization and age-, sex- and BMI-adjusted multivariable regression analyses for the associations between total sleep duration and 113 NMR-derived metabolomic measures. Each green dot in the scatter plot represents a metabolic trait and the positions of the dots are determined by the differences in mean metabolite concentrations (in standard deviation units) for each 1 hour greater reported total sleep duration. These are estimated by Inverse variance weighted (IVW) Mendelian randomization (vertical axes) and age, sex and BMI adjusted multivariable regression (horizontal axes). The vertical grey lines for each dot indicate the 95% confidence intervals (CI) for the Mendelian randomization estimates and the horizontal grey lines for each dot indicate the 95% CI for the adjusted multivariable regression estimates. A linear fit (red dashed line) summarizes the similarity between the two estimates. A slope of1 with an intercept of 0 (dashed grey line), with all green dots sitting on that line (R^2^ = 1), would indicate identical magnitude and direction between the two methods. R indicates goodness of linear fit and is a measure of the consistency between the two estimates. Abbreviations: AMV, adjusted (age, sex, BMI) multivariable regression; BMI, body mass index; CI, confidence interval; IVW MR., Inverse variance weighted Mendelian randomization, SD, standard deviation.

Associations for total sleep duration passed the multiple testing threshold for 8 of the 113 metabolomic trait associations in AMV analyses and one in MR. Only one of the 8 AMV associations replicated in the MR analyses (difference in mean creatinine for a 1 hour longer sleep was 0.02SD [0.01, 0.03] in AMV and 0.15 [0.02, 0.29] in MR). Isoleucine was the one metabolite to pass the multiple testing threshold in IVW MR analyses but it did not replicate in AMV analyses (0.01SD [−0.001, 0.02] in AMV and 0.22 [0.08, 0.36] in MR analyses]) (**Figure 6**). For the associations with creatinine, the weighted median MR result was consistent with that of the main (IVW) results but MR-Egger was in the opposite direction (though with very wide confidence intervals). For isoleucine, both MR sensitivity analyses had point estimates that were directionally, and in magnitude, similar to the main IVW MR results (**Figure 6**).

**Figure 6:**
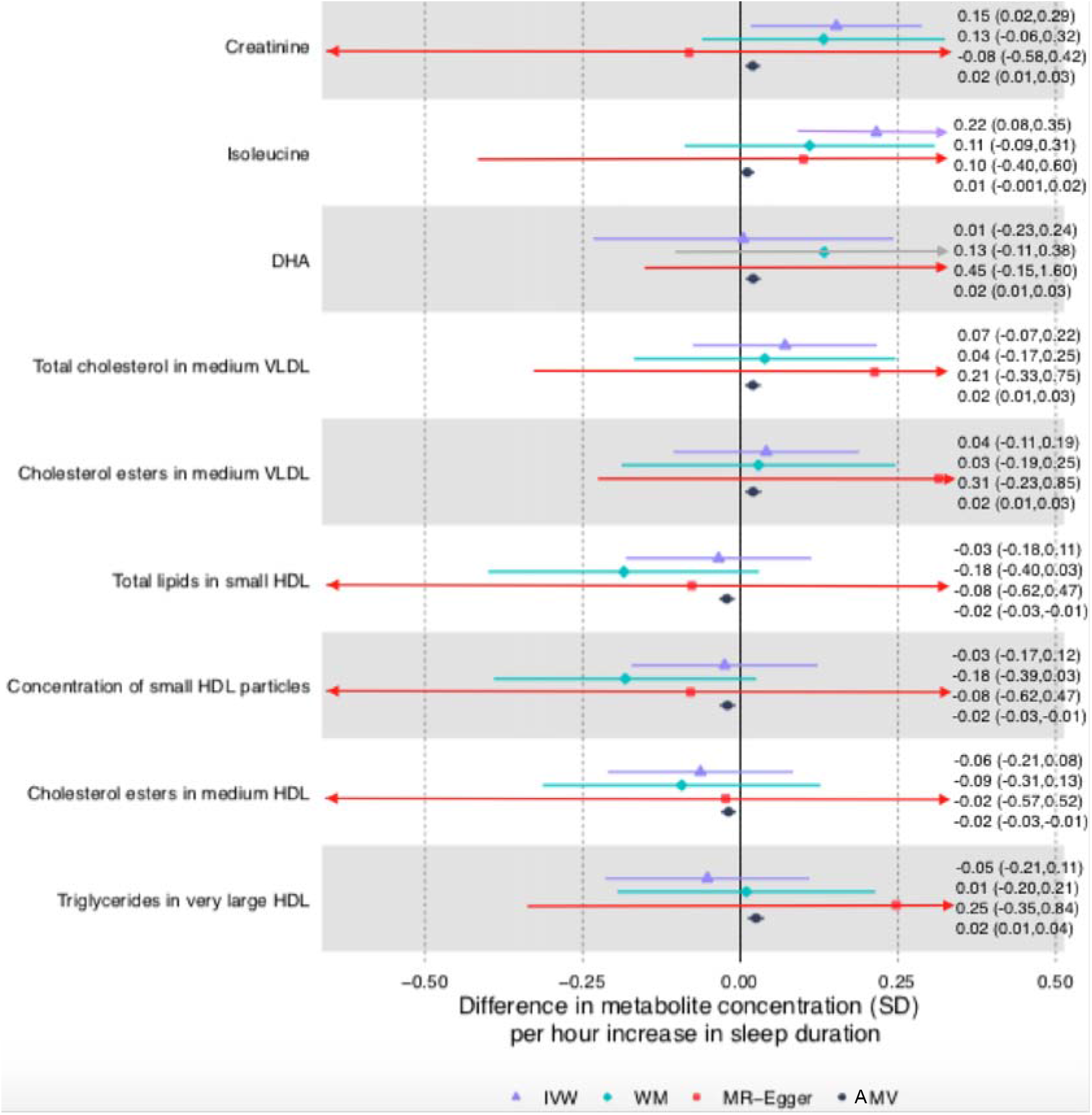
Mendelian randomization and age-, sex- and BMI-adjusted multivariable regression analyses results for selected associations of total sleep duration with NMR- derived metabolomic measures. Figure shows inverse variance weighted (IVW) Mendelian randomization, Mendelian randomization sensitivity (weighted median (WM) and MR-Egger) and adjusted multivariable (AMV) regression analysis results. Results presented were selected on the basis of passing multiple testing threshold for either IVW or AMV (p-values < 0.0029). The estimates are the difference in mean metabolite (in standard deviation units) per 1 hour greater total sleep duration. Abbreviations: AMV, adjusted (age, sex, BMI) multivariable regression; BMI, body mass index; DHA, 22:6, docosahexaenoic acid; HDL, high density lipoprotein; IVW MR, Inverse variance weighted Mendelian randomization; NMR, nuclear magnetic resonance; SD, Standard Error; WM, Weighted Median.

In exploratory analyses, most associations of short sleep duration (<7 hours) were close to the null in both AMV and MR analyses, with very little overall agreement between the two methods (R^2^=0.09, **Supplemental Figures S1** and **S2**). Two associations of short sleep passed the multiple testing corrected p-value in AMV analyses (22:6 docosahexonic acid (DHA) and omega-3 fatty acids), with short sleep duration associated with lower levels for both of these; none passed the multiple testing threshold in the MR analyses. For docosahexonic acid (DHA) and omega-3 fatty acids there was an inverse association in IVW MR analyses that had a larger effect estimate than in the AMV analyses but with wide confidence intervals that included the null (**Supplemental Figure S3**).

A total of 31 of the 113 metabolites passed the multiple testing threshold in the AMV analyses of long sleep duration (≥9 hours), including higher concentrations of most extremely large, large and medium VLDL, triglycerides, and concentrations of glycoprotein acetyls and isoleucine (**Supplemental Figures S4**). MR analyses did not support a causal effect for any of these, with MR analysis point estimate close to the null or in the opposite direction (**Figure S6**). We did not identify any metabolic traits passing the multiple testing threshold in IVW MR.

### Chronotype

There was very little consistency in direction and magnitude of association between AMV and MR analyses of chronotype with the metabolomic traits (**Figures 7 and 8**, R^2^ = 0.17). Chronotype was associated with isoleucine after multiple testing correction in the AMV analyses (difference in mean comparing evening to morning preference (0.13SD [0.04, 0.21]), but this was not supported in MR analyses (−0.02 [−0.05, 0.02])) (**Figure 9**). No associations of chronotype with the metabolomics traits passed the multiple testing threshold in the IVW MR analyses.

**Figure 7:**
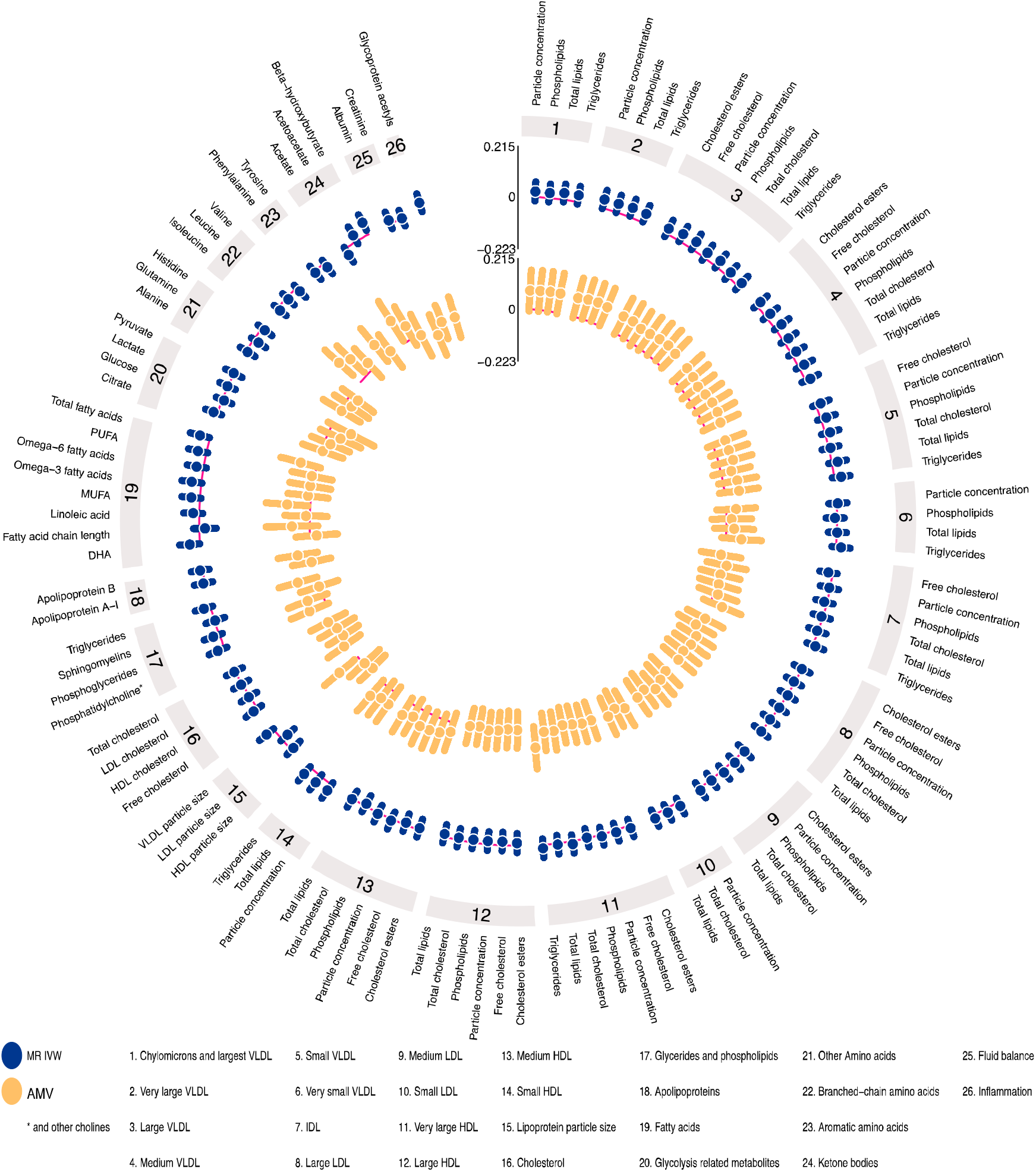
IVW Mendelian randomization estimates and age-, sex- and BMI-adjusted multivariable regression estimates for the associations between chronotype and 113 NMR-derived metabolomic measures. Results are the difference in mean metabolite concentrations (in standard deviation units) between those reporting an evening versus morning preference. Abbreviations: AMV, adjusted (age, sex, BMI) multivariable regression; BMI, body mass index; IDL, intermediate Results

**Figure 8:**
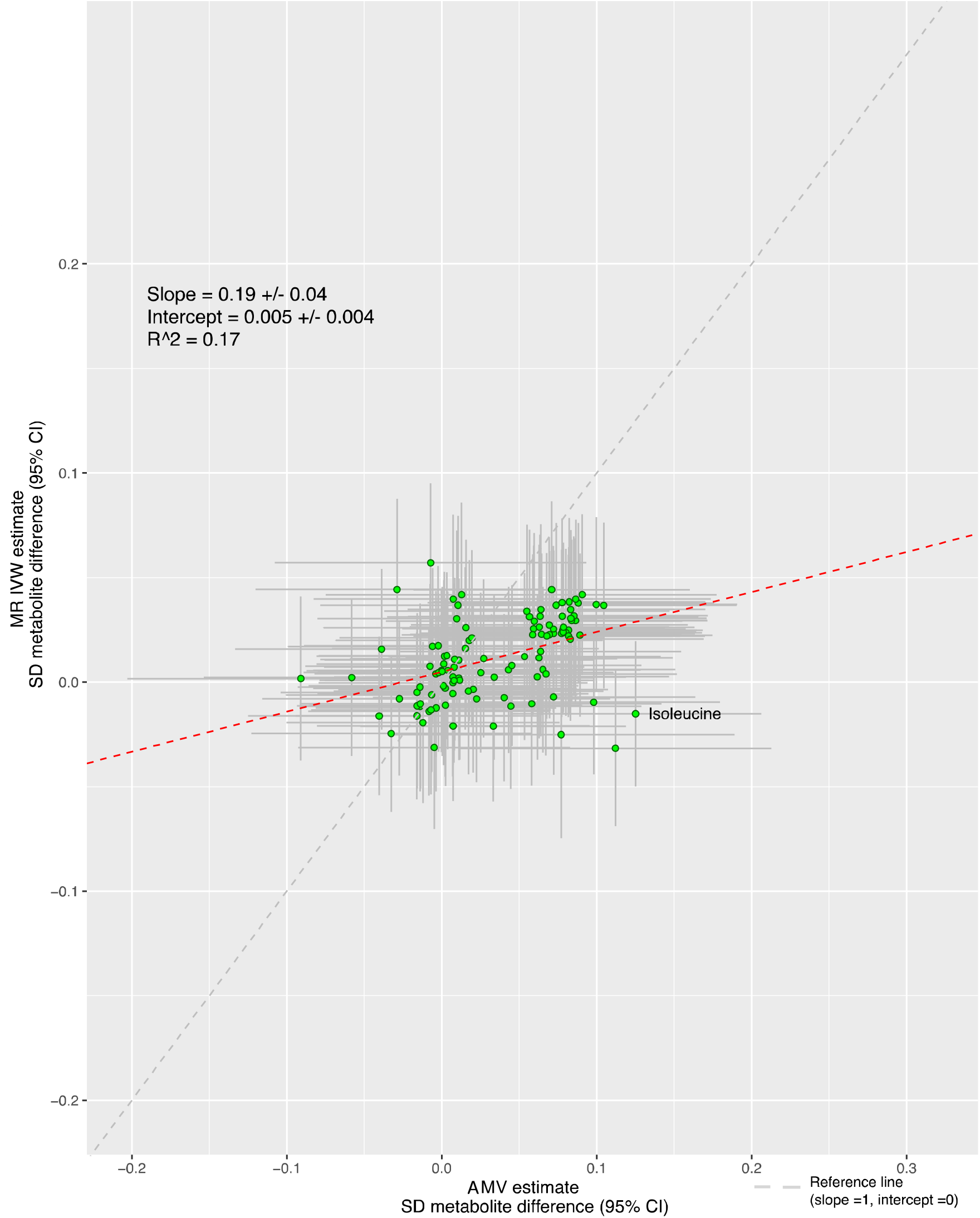
Comparison of the point estimates of the IVW Mendelian randomization and age-, sex- and BMI-adjusted multivariable regression analyses for the associations. Each green dot in the scatter plot represents a metabolic trait and the positions of the dots are determined by the differences in mean metabolite concentrations (in standard deviation units) comparing those reporting an evening preference versus morning preference. These are estimated by Inverse variance weighted (IVW) Mendelian randomization (vertical axes) and age, sex and BMI adjusted multivariable regression (horizontal axes). The vertical grey lines for each dot indicate the 95% confidence intervals (CI) for the Mendelian randomization estimates and the horizontal grey lines for each dot indicate the 95% CI for the adjusted multivariable regression estimates. A linear fit (red dashed line) summarizes the similarity between the two estimates. A slope of 1 with an intercept of 0 (dashedgrey line), with all green dots sitting on that line (R = 1), would indicate identical magnitude and direction between the two methods. R indicates goodness of linear fit and is a measure of the consistency between the two estimates. Abbreviations: AMV, adjusted (age, sex, BMI) multivariable regression; BMI, body mass index; CI, confidence interval; IVW MR, Inverse variance weighted Mendelian randomization, SD, standard deviation.

**Figure 9:**
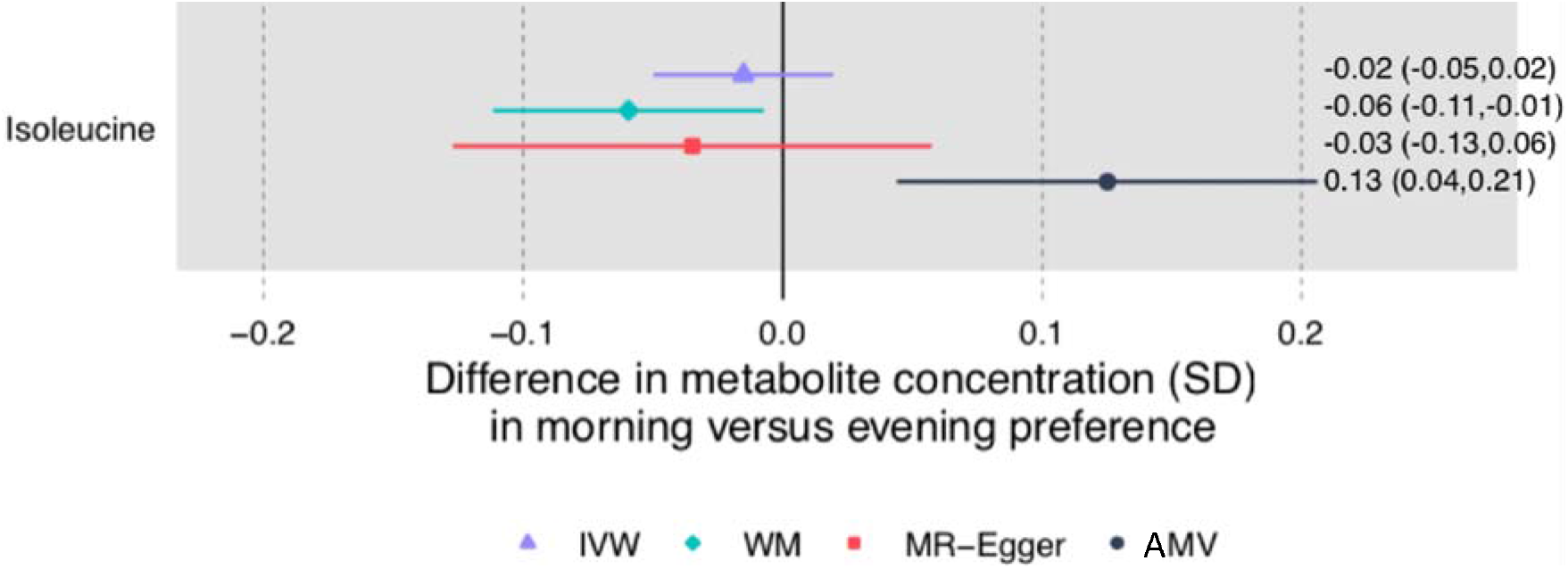
Mendelian randomization and age-, sex- and BMI-adjusted multivariable regression analyses results for selected associations of chronotype with NMR-derived metabolomic measures. Figure shows inverse variance weighted (IVW) Mendelian randomization, Mendelian randomization sensitivity (weighted median (WM) and MR-Egger) and adjusted multivariable (AMV) regression analysis results. Results presented were selected on the basis of passing multiple testing threshold for either IVW or AMV(p-values < 0.0029). The estimates are the difference in mean metabolite (in standard deviation units) comparing report of having an evening versus morning preference. Abbreviations: AMV, adjusted (age, sex, BMI) multivariable regression; BMI, body mass index; IVW MR, Inverse variance weighted Mendelian randomization; NMR., nuclear magnetic resonance; SD, standard deviation; WM, Weighted Median.

## Discussion

With the present multi-cohort effort, we intended to identify the potential biochemical mechanisms linking sleep to cardiometabolic disease risk. We found consistent evidence with both AMV and MR that usually (vs. sometimes, rarely or never) experiencing insomnia symptoms causes lower concentrations of citrate, total very large HDL particles and phospholipids in very large HDL particles and higher concentrations of glycoprotein acetyls. There was little consistency between AMV and MR results for total habitual sleep duration across all metabolomic traits, though a longer total sleep duration was associated with higher concentrations of creatinine in both methods. For chronotype, whilst having an evening preference was associated with higher isoleucine concentrations at our multiple-testing threshold in the AMV analyses, MR analyses did not support causality. Chronotype did not pass multiple testing with any other metabolites. Therefore, our findings do not support the notion that sleep traits have widespread effects on the investigated metabolomic traits. Nevertheless, they suggest that insomnia symptoms may influence cardiometabolic disease (as previously shown in MR^17^) through increased inflammation, and also result in lower citrate levels.

The lack of a more widespread impact of sleep traits on multiple metabolomic traits is in contrast with some experimental sleep studies, although direct comparisons are not possible. For example, targeted and untargeted mass spectrometry measurements performed in frequently sampled blood (every 2 hours) from 12 healthy men revealed that 109 out of 171 metabolites exhibited a circadian rhythm^43^. Furthermore, in controlled experimental conditions this circadian variation was maintained for 78 out of these 109 metabolites over a 24-hour period of total sleep deprivation. For 27 metabolites, including some lipids (13 glycerophospholipids and 3 sphingolipids), as well as tryptophan, serotonin, taurine and 8 acylcarnitines, marked acute increases in concentrations were observed during 24 hours of sleep deprivation compared with the 24 hours of habitual sleep^43^. Importantly, the MR analyses assessed long-term (lifelong), rather than acute, effects of a predisposition for unfavourable quality or quantity of sleep on metabolic disturbances, which could explain the generally stronger effects in the total sleep duration MR analyses.

Glycoprotein acetyls, which we identified as a novel trait potentially influenced by insomnia symptoms, are elevated in response to infection and inflammation. C-reactive protein (CRP) is the most widely recognized marker of acute and chronic inflammation in epidemiological studies. Whilst observational studies have shown that higher circulating CRP is associated with increased cardiovascular disease risk, MR studies suggest this is not a causal relationship^44, 45^. Glycoprotein acetyls have emerged as a potentially better measure of cumulative inflammation than CRP, since glycoprotein acetyls increase late in the inflammatory process and levels are relatively stable within individuals over many years^46, 47^. In AMV analyses in prospective cohorts, glycoprotein acetyls were positively associated with cardiovascular diseases and type 2 diabetes, independently of established risk factors and CRP^47^. If these associations are shown to be causal, then it is possible that cumulative chronic inflammation, as measured by glycoprotein acetyls, mediates the effect of insomnia on coronary heart disease identified in MR analyses^17^. However, we acknowledge that our results for the effect of insomnia on glycoprotein acetyls require replication in independent and larger studies and testing in ancestries other than Europeans.

The inverse association of insomnia symptoms with citrate in both AMV and MR analyses is novel. A recent narrative review highlighted the physiological control of plasma citrate concentrations in health and disease^48^. One possible mechanism through which insomnia might influence citrate is via the relationship of insomnia with night-time eating (which is also accompanied with higher night physical activity)^49^, which would result in higher TCA cycle activity and consequently lower plasma citrate concentrations. However despite a plausible role, there is a paucity of clinical and epidemiological studies of the effect of citrate levels on disease outcomes^48^. Therefore, the meaning of a possible effect of insomnia on citrate levels is hard to discern.

We found evidence for associations of experiencing insomnia symptoms with higher concentrations of very large total HDL particles and phospholipids in very large HDL particles. MR and randomized controlled trials suggest that circulating HDL cholesterol is not causally related to cardiovascular disease^50-52^. The amount of cholesterol carried in HDL particles increases with increasing particle size and emerging evidence highlights the importance of considering size, structure and composition of lipoprotein particles when exploring their effects on cardiovascular disease ^53^. In AMV analyses, inverse associations of very large, large, medium and small HDL particles with cardiovascular disease have been observed, but these attenuated to the null with adjustment for lipids used by clinicians^36^. Thus, the relevance of possible insomnia effects on very large HDL particle concentrations, and specifically phospholipids in these particles, is unclear and require additional studies.

We found evidence in both AMV and MR analyses of a possible association of longer total sleep duration with higher creatine concentrations, a biomarker used to estimate kidney function. Established cardiovascular risk factors, such as high blood pressure and type 2 diabetes, are associated with higher creatinine concentrations^54^. Findings from multivariable regression suggest that the association of kidney function with cardiovascular disease largely reflects confounding and/or reverse causality^55^. Thus, our observations possibly suggest that longer sleep duration is an additional risk factor for chronic kidney disease rather than cardiovascular diseases, though we acknowledge MR sensitivity analyses did not support a causal effect. We also found a novel association of longer total sleep duration with the branched-chain amino acid isoleucine in MR analyses. Higher concentrations of branched-chain amino acids, including isoleucine, are associated with increased risk of cardiovascular disease^36^, though this has not been explored in MR studies. MR analyses supports a causal effect of the branched-chain amino acids on type 2 diabetes^56^ and our results suggest that longer total sleep duration may mediate some of this effect.

Key strengths of our study are its novelty and the comparison of results from the largest AMV study of sleep traits with multiple circulating metabolomic measures^22^ with equivalent results from MR. We harmonised questionnaire-based sleep data across all contributing studies and the NMR metabolomic platform was consistent across studies in both the AMV and MR analyses. We were able to increase the power of our two-sample MR analyses by combining unpublished summary-level GWAS results from three cohorts (total N=13,693) with those of the largest published GWAS of the same NMR platform (N=24,925) to date^31^. Two-sample MR assumes that the two samples are from the same underlying population and independent of each other. Given all GWAS were undertaken in adults of European ancestry and the lack of overlap in studies contributing to the metabolite GWAS with any of the sleep trait GWAS, we are confident this assumption is largely met. Most observed differences in mean metabolomic concentrations were close to the null, and in general (true) null results are less subject to bias than non-null results^57^.

Important limitations include the lack of statistical power, particularly to explore possible non-linear associations for sleep duration. The platform misses a high proportion of currently quantifiable metabolites in human serum/plasma, including markers of energy balance, microbiota metabolism, vitamins, co-factors and xenobiotics, that may be influenced by sleep traits^43^. Still the NMR platform used in the analyses covers considerably more of the lipidome than conventional clinical chemistry measures (total cholesterol, LDL-C, HDL-C and triglycerides) that have previously been explored, and in addition includes amino acids, glycolysis metabolites, ketone bodies and an inflammatory marker. Whilst we adjusted for age, sex and BMI, the results obtained in multivariable-adjusted regression may be exaggerated by residual confounding from unobserved confounders such as socioeconomic position, smoking, and physical activity. As the AMV results were cross-sectional it is also possible that variation in metabolomic traits influences sleep patterns, and some of the multivariable regression results not verified in MR are due to reverse causality. In addition, we restricted the analyses to cohorts containing mostly European participants (one cohort contributing to AMV meta-analysis, HELIUS, included non-European participants, whereas all MR analyses were restricted to Europeans). This reduces the potential for population stratification to bias our MR analyses, but hampers generalization of our findings to other ancestry groups. The MR results may have been influenced by weak instrument bias, which, if present, would be expected to bias results towards the null. Sensitivity analyses exploring possible bias due to directional horizontal pleiotropy were mostly consistent with the main IVW findings, though MR-Egger estimates, were imprecise as expected with this method which is statistically less efficient than the main IVW method.

Taken together, our findings do not suggest widespread metabolic disruption caused by sleep traits. However, the evidence for possible effects of insomnia symptoms on glycoprotein acetyls and citrate, and longer total sleep duration on creatinine and isoleucine might explain some of the effects, found in MR analyses, of these sleep traits on cardiometabolic diseases. These warrant further investigation.

## Data Availability

RN and DAL act as guarantors for the paper.

## Acknowledgements

We are most grateful to the participants of the different participating studies. In addition, we are grateful to the management team, research nurses, interviewers, research assistants and other staff who have taken part in gathering the data of the different studies.

